# Gene regulation contributes to explain the impact of early life socioeconomic disadvantage on adult inflammatory levels in two European cohort studies

**DOI:** 10.1101/2020.04.03.20050872

**Authors:** Cristian Carmeli, Zoltán Kutalik, Pashupati P. Mishra, Eleonora Porcu, Cyrille Delpierre, Olivier Delaneau, Michelle Kelly-Irving, Murielle Bochud, Nasser A. Dhayat, Belen Ponte, Menno Pruijm, Georg Ehret, Mika Kähönen, Terho Lehtimäki, Olli T. Raitakari, Paolo Vineis, Mika Kivimäki, Marc Chadeau-Hyam, Emmanouil Dermitzakis, Nicolas Vuilleumier, Silvia Stringhini

## Abstract

Individuals growing up during childhood in a socioeconomically disadvantaged family experience a higher rate of inflammation-related diseases later in life. Little is known about the mechanisms linking early life experiences to the functioning of the immune system decades later. Here we explore the relationship across social-to-biological layers of early life social exposures on levels of adulthood inflammation (C-reactive protein) and the mediating role of gene regulatory mechanisms, epigenetic and transcriptomic profiling from blood, in 2,329 individuals from two European cohort studies. Consistently across both studies, we find transcriptional activity explains a substantive proportion (up to 78%) of the estimated effect of early life disadvantaged social exposures on levels of adulthood inflammation. Furthermore, we show that mechanisms other than DNA methylation potentially regulate those transcriptional fingerprints. These results further our understanding of social-to-biological transitions by pinpointing the role of pro-inflammatory genes regulation that cannot fully be explained by differential DNA methylation.

## INTRODUCTION

Inflammatory processes have been suggested as a potential pathophysiological mechanism underlying the development of several major chronic diseases, including cardiovascular, metabolic and psychotic disorders^1,2,3^. Systemic inflammation is also one of the most studied pathways in the context of the social-to-biological transition^4,5,6,7,8^. Although early life disadvantaged socioeconomic conditions have been consistently shown to lead to elevated levels of inflammation in adulthood^9^, the underlying biological processes through which those socioeconomic exposures are embodied remain unclear.

Various studies have suggested a dysregulation of pro-inflammatory genes transcription in leukocytes as a potential biological mechanism for the embodiment of socioeconomic disadvantage^10,11,12,13,14^. This line of reaserch rests on the hypothesis that socioeconomic disadvantage activates the sympathetic nervous system associated to body’s flight or fight response to stressors that eventually leave a leukocyte transcriptional fingerprint. Similar findings have been reported in experiments conducted with macaques^15,16^. Other studies have focused on the epigenetic regulators of transcriptional activity in leukocytes, in particular the DNA methylation levels at specific CpG sites^17,15,18^. These research endeavors were conducted without directly relating DNA methylation to gene transcription levels, whose functional relationship does not always hold^19,20^.

To date, the contribution of pro-inflammatory genes regulation to socioeconomic inequalities in inflammatory levels has not been investigated empirically, although frequently discussed theoretically^10,17^. To explore the chain of biological mechanisms linking early life socioeconomic disadvantage to adult inflammatory levels via DNA methylation and gene transcription in leukocytes, we used data from a Finnish (1,623 participants) and a Swiss (706 participants) population-based cohort study, and evaluated the consistency of findings across these two populations. First, we calculated the portion of the effect of early life socioeconomic disadvantage on adult heightened inflammatory levels explained by transcriptional level of pro-inflammatory genes either inferred by applying 2-sample Mendelian randomization^21^ on the whole transcriptome (2sMR) or belonging to the conserved transcriptional response to adversity (CTRA) model^12^. We measured levels of systemic inflammation through circulating C-reactive protein (CRP), as it is a sensitive marker of inflammation and elevated amounts have been associated with increased risk for several diseases^3^. Second, we established the most likely functional relationship between *cis* (based on their genomic location) epigenetic and transcriptional changes induced by early life socioeconomic disadvantage via Bayesian networks model selection^22^.

## RESULTS

### Pro-inflammatory genes selection via 2sMR

We used summary statistics of the associations between genetic instruments (Single Nucleotide Polymorphisms, SNPs) and gene transcription in leukocytes or CRP in blood obtained via large-scale GWAS conducted by the eQTLGen consortium^23^ and the CHARGE consortium^3^, respectively (see Methods). There were 10,701 genes for which 2sMR provided a test of the causal association between gene transcription and CRP levels. There was an average of 39 semi-independent instruments (*cis* eQTLs in a linkage disequilibrium r^2^≤0.25, see Methods) per each gene and the average *F*-statistic was 97. Finally, 81 genes were significant at *q*≤0.05 (see Table S1 and Methods). Of note, one of them, *NFKB1*, was as well one of the CTRA genes. Functional enrichment analysis revealed enriched inflammatory bowel disease, influenza A and prion disease pathways.

### Characteristics of populations

As reported in Table 1, on average Finnish participants (YFS study) were 42-year old (age ranging between 34 and 49 years), while Swiss participants (SKIPOGH study) were 53-year old (age ranging between 25 and 88 years). Women represented 46% of the YFS population and 52% of the SKIPOGH population. CRP levels in SKIPOGH were distributed differently than in YFS, as participants in the upper quintile had CRP≥3 mg/L in SKIPOGH while had CRP>2.2 mg/L in YFS (see Table 1). Individuals with high/low parental occupational position were 18%/26% in YFS and 25%/28% in SKIPOGH.

**Table 1.** Characteristics of the YFS and SKIPOGH populations. Participants are those with complete information about parental occupational position, transcriptome and CRP. Categorical characteristics are summarized through their absolute and relative (%) prevalence. Age is summarized through its mean and range. CRP in YFS is summarized with median and lower to upper quintiles range, while in SKIPOGH is binarized as 3mg/L is the highest left truncation value (see Methods).

| Characteristics | YFS | SKIPOGH |
| --- | --- | --- |
| N | 1,623 | 706 |
| Parental occupational position<br>[high/intermediate/low] | 289 (17.8%)<br>907 (55.9%)<br>427 (26.3%) | 176 (24.9%)<br>329 (46.6%)<br>201 (28.5%) |
| Parental education<br>[high/intermediate/low] | 439 (30.9%)<br>480 (33.7%)<br>504 (35.4%) | 187 (26.5%)<br>216 (30.6%)<br>303 (42.9%) |
| Center | 238 (14.7%)<br>343 (21.1%)<br>278 (17.1%)<br>503 (31.0%)<br>261 (16.1%) | 106 (15.0%)<br>290 (41.1%)<br>310 (43.9%) |
| Sex [man/woman] | 882 (54.3%)<br>741 (45.7%) | 341 (48.3%)<br>365 (51.7%) |
| Age [years] | 42 [34,49] | 53 [25,88] |
| CRP [mg/L] | 0.7 [0.3,2.2] | CRP<3mg/L: 560 (79.3%)<br>CRP≥3mg/L: 146 (20.7%) |

### Mediation by genes transcription

Individuals with low parental occupational position (potentially counter to fact) had an increase in (the geometric mean of) adulthood CRP of 22.6% [95% confidence intervals (CI): 2.7%, 46.4%] for YFS and 65.1% [95% CI: 32.7%,106.9%] for SKIPOGH compared to having (potentially counter to fact) high parental occupational position (total effect, see Figure 1). The point estimate of the geometric mean of CRP for individuals with low parental occupational position was 0.75 mg/L in YFS and 0.52 mg/L in SKIPOGH, while for individuals with high parental occupational position was 0.61 mg/L in YFS and 0.32 mg/L in SKIPOGH.

**Figure 1.**
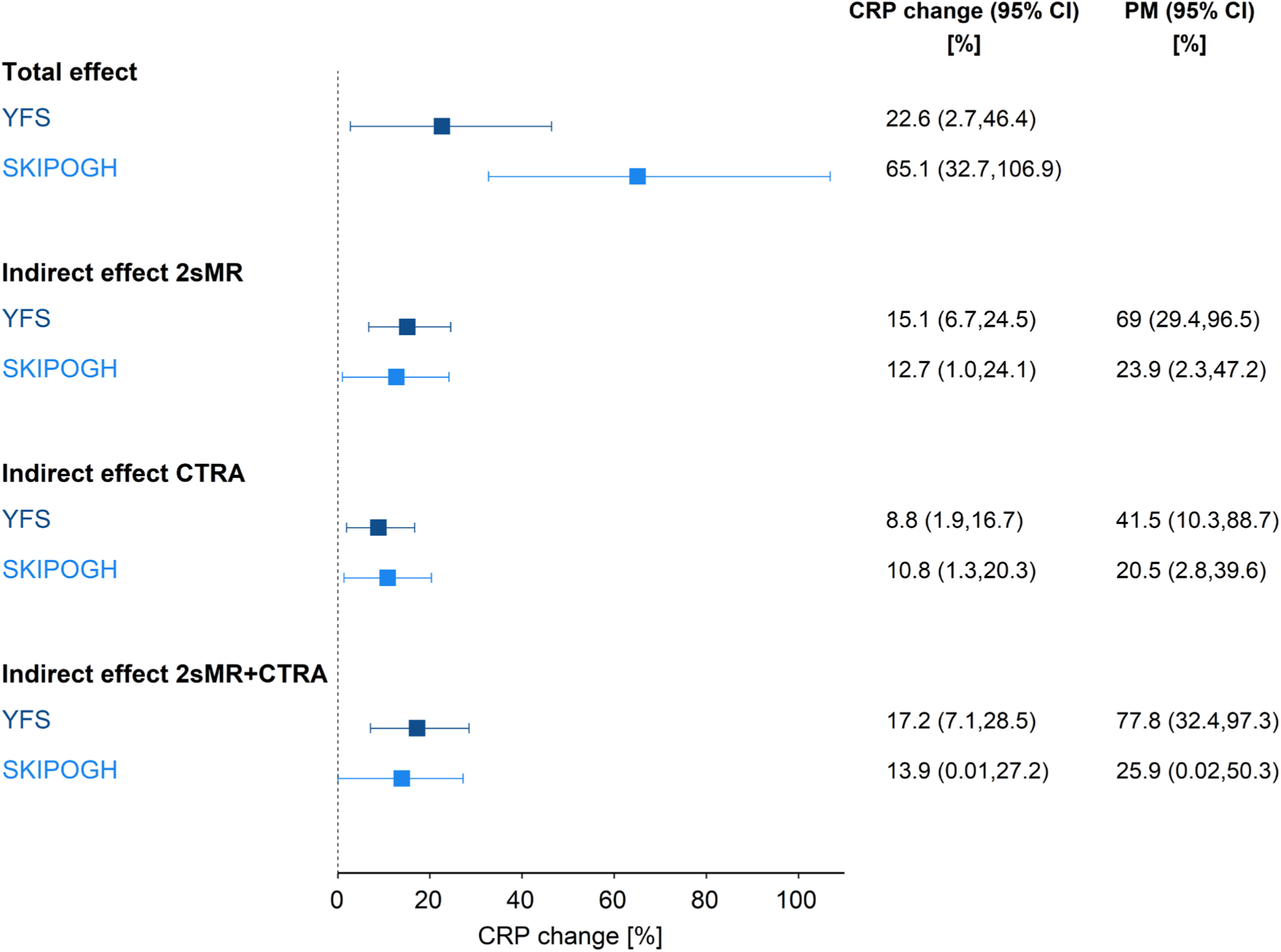
Size of effects and proportion mediated (PM) estimated by mediation analysis in each population. CRP change (%) and 95% confidence intervals (CI) are reported for the total effect of low (vs high) parental occupational position on CRP levels in adulthood, and for indirect effects through transcription levels of pro-inflammatory genes *en-bloc*. Joint indirect effects via 2sMR, CTRA or 2sMR+CTRA genes were estimated by summarizing transcription levels with principal components corresponding to about 50% of the transcription variance in each genes’ set.

The portion of the total effect accounted by 2sMR-implicated genes *en-bloc* (indirect effect, whereby 2sMR genes transcription levels were summarized by the first 23 principal components (PCs) corresponding to 49.9% of transcription variance, see Methods) resulted in an increased (geometric mean of) CRP of 15.1% [95% CI: 6.7%, 24.5%] in YFS, and of 12.7% [95% CI: 1.0%,24.1%] in SKIPOGH (first 23 PCs corresponding to 50.4% of transcription variance). This corresponded to a proportion mediated (PM, see Methods) of 69.0% [95% CI: 29.4%, 96.5%] in YFS and of 23.9% [95% CI: 2.3%, 47.2%] in SKIPOGH. The indirect effect accounted by CTRA genes *en-bloc* (first 12 PCs corresponding to 49.3% of transcription variance) resulted in an increased (geometric mean of) CRP of 8.8% [95% CI: 1.9%,16.7%] in YFS, and of 10.8% [95% CI: 1.3%,20.2%] in SKIPOGH (first 10 PCs corresponding to 49.8% of transcription variance). The corresponding PM was 41.5% [95% CI: 10.3%, 88.7%] in YFS and 20.5% [95% CI: 2.8%, 39.6%] in SKIPOGH. When considering 2sMR-implicated and CTRA genes together (133 in total, see Table S1), their joint indirect effect resulted in an increased (geometric mean of) CRP of 17.2% [95% CI: 7.1%,28.5%] in YFS (first 33 PCs corresponding to 50.1% of transcription variance) and of 13.9% [95% CI: 0.0%,27.2%] in SKIPOGH (first 30 PCs corresponding to 49.8% of transcription variance). This corresponded to a PM of 77.8% [95% CI: 32.4%, 97.3%] in YFS and of 25.9% [95% CI: 0.02%, 50.3%] in SKIPOGH. The partial redundancy between the two gene sets was supported by correlation patterns between the principal components and between the transcription levels (see Figures S1 and S2).

Overall, in both cohorts the magnitude of effects was in the same direction and that of indirect effects was similar, with estimates of 17.2% and 13.9% indicating that pro-inflammatory genes transcription mediates a substantive portion of the estimated total effect of parental occupational position on adult CRP.

### Regulation of genes transcription by local DNA methylation

We applied Bayesian network scoring to 4,076 CpGs located nearby (<1500 bp) the 2sMR-implicated and CTRA genes (see Table S2), for five causal structures (Group A to E, see Figure S5 and Methods). As described in Table 2, for most CpGs (Group A: N=3,612 (88.6%) and N=3,253 (79.8%) in YFS and SKIPOGH, respectively), there was no substantive evidence (logarithm of Bayes factor larger than one, see Methods) for an influence of parental occupational position on adult regulation of genes transcription. For one CpG (cg19527571), there was evidence for the role of DNA methylation as *cis*-regulator of *MED24* in SKIPOGH only (Group B). There was evidence for parental occupational position driving levels of gene’s transcription and DNA methylation being either unrelated to transcription (Group C: N=30 and N=36 CpGs in YFS and SKIPOGH, respectively) or driven by transcription (Group D: N=11 and N=17 CpGs in YFS and SKIPOGH, respectively). For 9 and 24 CpGs (Group E, YFS and SKIPOGH, respectively) there was evidence for parental occupational position driving levels of methylation but not transcription levels. Finally, there was no substantive evidence for one causal structure over the others in N=414 (10.2%) and N=745 (18.3%) CpGs in YFS and SKIPOGH, respectively.

**Table 2.** Cardinality of the most likely group of model structures across the five tested competing groups (see Figure S5) for a total of 4,076 CpG sites in up to N=1,367 YFS and N=661 SKIPOGH participants. The most likely group of causal structures is selected when its posterior probability is at least roughly three times larger than any other group’s posterior probability. None corresponds to CpGs for which none of the five tested groups reached winning evidence or models could not be estimated as the transcription level was not available in a few genes (37 and 33 CpGs in YFS and SKIPOGH, respectively).

| Causal structures |  | N of CpGs |  |
| --- | --- | --- | --- |
|  |  | YFS | SKIPOGH |
| A | Parental occupational position does not influence methylation nor transcription levels | 3,612 | 3,253 |
| B | Parental occupational position drives changes in methylation and transcription levels; methylation also causes changes in transcription | 0 | 1 |
| C | Parental occupational position drives gene transcription, but methylation and transcription are not related | 30 | 36 |
| D | Parental occupational position drives transcription that in turn influences methylation levels | 11 | 17 |
| E | Parental occupational position influences methylation but not transcription levels | 9 | 24 |
| None |  | 414 | 745 |
| Total |  | 4,076 | 4,076 |

Overall, a non-empty set of CpGs was present across the two cohort studies for Groups A, C, D and E. Furthermore, this consistency at pattern level did correspond to a replication at the level of single site for 2,894 CpGs belonging to Group A only (see Table S2).

Compared to CpGs in Group A, sites in either Group C or D (N=41 and N=53 in CRYFS and SKIPOGH, respectively) had similar distribution in CpG island position (island or shore or shelf vs open sea, χ^2^=0.1 and 0.2, *P* value=0.76 and 0.48, %, YFS and SKIPOGH, respectively) and increased TSS or UTR annotation (56% vs 44% and 64% vs 44%, χ^2^=2.8 and 7.9, *P* value=0.15 and 0.005, YFS and SKIPOGH, respectively).

### Sensitivity analyses

Total and indirect effect estimates were in the same direction and of similar (for YFS) and higher (for SKIPOGH) magnitude to those reported in main analysis when excluding participants with high values of CRP (>10 mg/L, N=34 [2.1%] in YFS and N=12 [1.7%] in SKIPOGH) (see Figure S3). By varying the amount of variance explained by principal components summarizing transcription levels of the genes’ sets, indirect effect estimates slightly increased when variance moved from 30% to 50% and remained stable when variance moved from 50% to 60% (see Table S3). This finding supports the fact that indirect effects reported in main analyses do not underestimate the amount of mediation by pro-inflammatory genes regulation. In SKIPOGH, when using a dichotomous CRP (≥3 mg/L) as outcome, the estimated indirect effect was substantive (see Table S4), backing the results presented in main analyses with an imputed continuous CRP.

When parental education was considered as the exposure, the magnitude of indirect effects was in the same direction and slightly lower than that reported in main analysis (see Figure S4). Furthermore, the Bayesian scoring procedure supported a pattern of models similar to the one found for parental occupational position, but no consistent evidence for models whereby parental education drives transcription which then affects DNA methylation levels (see Table S5). When adding a group of negative control DAGs to the groups described in Figure S5, the Bayesian scoring procedure correctly did not assign that group to any of the CpGs.

## DISCUSSION

We found that both Finnish and Swiss adults having experienced disadvantaged socioeconomic conditions early in life displayed heightened levels of systemic inflammation compared to their more advantaged counterparts, and that a substantive portion of this effect was transmitted by shifts in leukocytes pro-inflammatory genes transcription. Via 2sMR, we identified a new set of pro-inflammatory genes putatively influenced by socioeconomic conditions in early life. Finally, we showed a consistent pattern of inter-individual variation in local DNA methylation being either unrelated to or driven by inter-individual variation in transcription.

To our knowledge, this is the first study examining regulatory mechanisms linking early life socioeconomic conditions to inflammatory levels in adulthood by integrating epigenetic and transcriptomic profiling from blood in two European populations. The observed socioeconomic inequalities in CRP (that is the estimated total effect) were in line with a previous multi-cohort study (N=13,078) from four European countries^6^, whereby the pooled association between paternal occupational position and CRP resulted in an increased (geometric mean of) CRP of 20.9% [95% CI: 11.6%, 31.0%]. In our study, some differences across the two cohorts were observed in the estimated magnitude of total and indirect effects. As populations were sampled using different designs, were from different European countries and had a different age distribution, these characteristics may have influenced those inter-cohort differences. Remarkably, in both populations those effects were in the same direction and the magnitudes of mediation were similar. Indeed, the joint indirect effect estimates were 17.2% and 13.9% (2sMR+CTRA genes in YFS and SKIPOGH, respectively), indicating a substantive mediation. However, the magnitude of the indirect effects can be an overestimate since unmeasured confounding may have increased the association between genes transcription and heightened inflammation. Furthermore, another limitation of our study is that CRP was not measured through a high-sensitivity assay in SKIPOGH. However, main and sensitivity analysis provided consistent results, and two thirds of SKIPOGH participants had CRP measured with high precision at 2 mg/L (coefficient of variation<5%). Overall, our finding of a mediating role of pro-inflammatory genes regulation is supported by previous studies in humans and primates. Those research endeavours supported transcriptional fingerprint of decreased glucocorticoid and increased pro-inflammatory signalling as key mechanism in the embodiment of socioeconomic disadvantage^10,15^.

Our transcriptome-wide analysis also identified 81 pro-inflammatory genes via 2sMR, a method that is protected from confounding and reverse causation biases. By using summary statistics from two large data sets we increased statistical power, and by performing inference via three 2sMR methods based on orthogonal assumptions we strengthened the reliability of inference. Nevertheless, we cannot exclude bias in our 2sMR analysis due to selection of instruments in the same data set where we conducted inference^25^, which leads to underestimation of causal effect sizes. Furthermore, we applied a stringent, rather conservative threshold for genes’ selection, and we could not apply 2sMR to all human genes, therefore we cannot exclude the existence of other undetected pro-inflammatory genes.

We showed in both Finnish and Swiss individuals that early in life socioeconomic conditions may regulate gene transcription in leukocytes without involving local DNA methylation changes, or with local DNA methylation playing the role of regulated event. This finding is in line with recent experimental observations obtained via a novel sequencing technology^19^, Bayesian network analyses of ancestry-related differences in DNA methylation/gene transcription in cord blood^35^ and whole blood monocytes^36^, and a Mendelian randomization study^24^. As pointed out therein^24^, the inferred direction of causality between DNA methylation and gene transcription can be biased because their measurement is noisy. Nevertheless, our findings are based on interpreting only models with substantial statistical evidence (logarithm of Bayesian factor larger than one). At the same time, that threshold could have biased our findings, as we could not assign any of the investigated causal structures to about 10% of the CpGs. Our results do not exclude the possibility that at some *cis* CpGs methylation levels do drive gene transcription and they point to the presence of other regulating mechanisms. Overall, this finding calls attention to potential weaknesses of current epidemiologic approaches based on investigating a single epigenetic layer uncoupled with downstream functional molecular phenotypes.

Our study supports regulation of pro-inflammatory genes in leukocytes as a mechanism through which disadvantaged early life socioeconomic conditions heightens inflammatory levels in adult life, but it is still unclear when that happens and via which intermediate exposure. Repeated measurements across the life course are needed to answer that question. Our work could extend in prioritizing repeated measurements in life course longitudinal studies to identify critical/sensitive windows of socioeconomic exposures.

## METHODS

### Study populations

We used individual-participant data from two multi-centre population-based studies: the Cardiovascular Risk in Young Finns Study (YFS)^26^ and the Swiss Kidney Project on Genes in Hypertension (SKIPOGH)^27^.

YFS participants were selected from five Finnish areas according to the location of the university cities with a medical school (Helsinki, Kuopio, Oulu, Tampere and Turku). In each area, urban and rural boys and girls were randomly selected on the basis of their personal social security number from the Social Insurance Institution’s population register, which covers the whole Finnish population. The baseline survey was conducted in six age cohorts of children (aged 3, 6 and 9) and adolescents (aged 12, 15 and 18) in 1980, with a participation rate of 83%, i.e. 3,596 of those invited. Participants were re-examined every 3 years between 1980 and 1992, in 2001 and in 2011, when they had reached mid adulthood (aged 34–49). The YFS study was conducted according to the guidelines of the Declaration of Helsinki, and the protocol was approved by local ethics committees. Own/parental consent was obtained for all participants.

SKIPOGH participants were recruited in the cantons of Bern and Geneva and the city of Lausanne, Switzerland. Participants were randomly selected from the population-based CoLaus study in Lausanne^28^, and from the population-based Bus Santé study in Geneva^29^. In Bern, participants were randomly selected using the cantonal phone directory. Inclusion criteria were: (1) written informed consent; (2) minimum age of 18 years; (3) white ethnicity; (4) at least one, and preferably three, first-degree family members also willing to participate. Participation rate was 20% in Lausanne, 22% in Geneva, and 21% in Bern. The baseline assessment initiated in December 2009 and ended in April 2013, including 1,128 participants aged between 18 and 90 years. A follow-up survey (1,033 individuals) started in October 2012 and was completed in December 2016. The SKIPOGH study was approved by the ethical committees of Lausanne University Hospital, Geneva University Hospitals and Bern University Hospital.

### Causal models and measures

The causal structures posited in our study are represented in the directed acyclic graphs (DAGs) displayed in Figure 2. The DAG in figure 2A evaluates the portion of the effect of early life socioeconomic conditions (exposure) on adult inflammatory levels (outcome) transmitted by intracellular processes regulating transcription of genes in immune cells (mediators). This “indirect” effect captures all potential pre- and post-transcriptional regulating mechanisms including DNA methylation, histone modifications, transcription factors binding and microRNAs^30,31^. These processes mutually regulate in both physiological and diseases conditions. In the second part of our study, we focused on local DNA methylation as regulating mechanism (mediator) of a gene’ transcription (outcome) according to the causal structure in Figure 2B. We chose DNA methylation as it is the most widely studied epigenetic mechanism in the field of social epidemiology^32^ and as it represents a potential therapeutic target^33^. Since the relationship between levels of local DNA methylation and genes transcription can be bidirectional^34,35,36^ or absent^19^, for each pro-inflammatory gene and DNA methylation site we did not posit any specific causal structure between gene transcrition and DNA methylation measurements. Instead, we investigated the most likely one among all possible causal structures (see Figure S5 and section entitled *Bayesian network model selection*).

**Figure 2.**
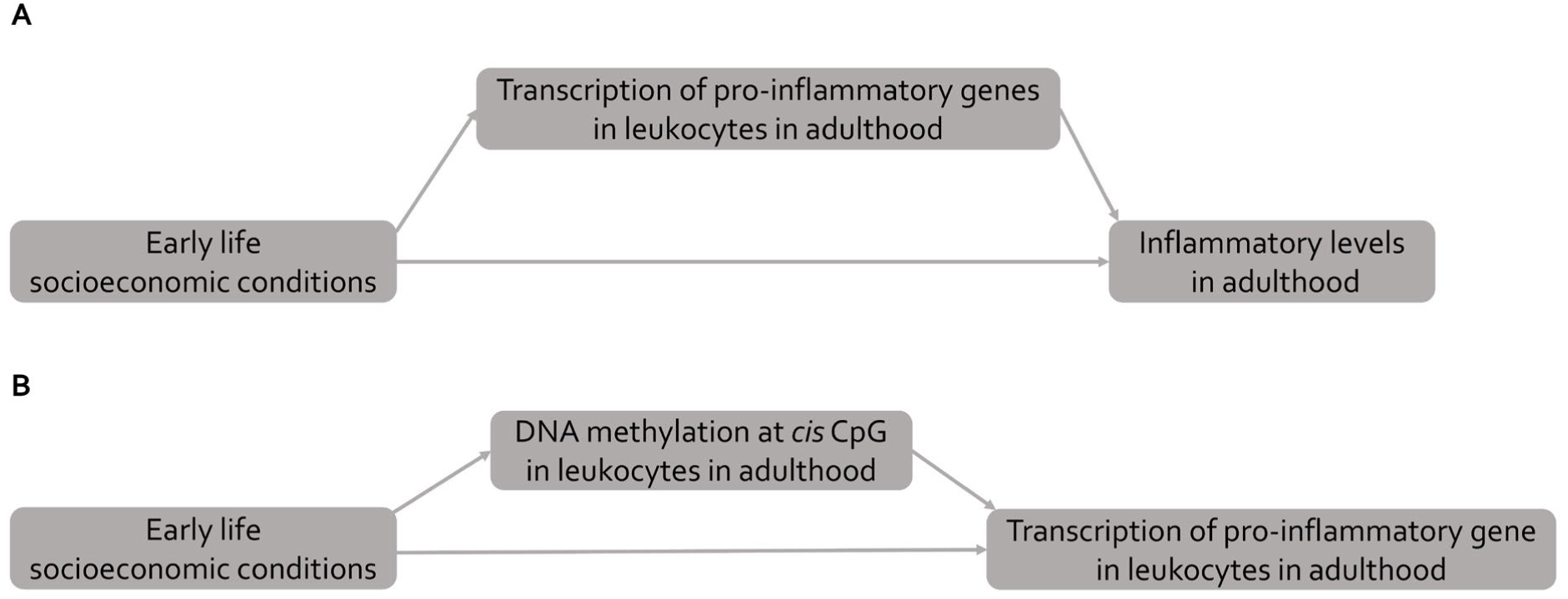
Causal structures posited in our study. For the sake of simplicity, we do not draw confounders (age, sex, centre of blood collection). Early life socioeconomic conditions represent the exposure. A) Inflammatory levels in adulthood represents the outcome, and transcription of pro-inflammatory leukocytes genes in adulthood the investigated intermediate mechanism. B) Transcription level of a pro-inflammatory gene in leukocytes represents the outcome and DNA methylation levels at a specific *cis* CpG in leukocytes the mediating mechanism.

Parental occupational position defined as the highest household occupation was used as an indicator of socioeconomic conditions in early life (main analysis). In YFS, the occupational position (manual, lower-grade non-manual and higher-grade non-manual, farmer) of the head of the household was assessed in 1980 (study’s baseline), while SKIPOGH participants reported at the study’s follow-up the profession of their father when they were children. Paternal occupational position is a commonly used indicator of socioeconomic conditions in early life^37^ and is closely related to parental occupational position in the Swiss context until the early 1980s (see SI Appendix). In a sensitivity analysis, we adopted a measure of parental education as an alternative proxy of socioeconomic conditions in early life (see SI Appendix).

Occupational position was categorized according to the European SocioEconomic Classification (ESEC) scheme into high (higher professionals and managers, higher clerical, services, and sales workers (European socioeconomic class 1-3)), intermediate (small employers and self-employed, farmers, lower supervisors and technicians, class 4-6), or low (lower clerical, services, and sales workers, skilled workers, semi-skilled and unskilled workers, class 7-9)^38,39^.

Systemic inflammatory status was assessed through the amount of C-reactive protein (CRP [mg/L]) in serum (YFS) and plasma (SKIPOGH). CRP is a sensitive marker of inflammation and elevated amounts of CRP have been associated with increased risk for several diseases^3^. In YFS, CRP levels were measured from blood collected in 2011 through an automated analyzer (Olympus AU400; Tokyo, Japan) and a highly sensitive turbidimetric immunoassay kit (‘CRP-UL’-assay; Wako Chemicals, Neuss, Germany). Detection limit of the assay was 0.06 mg/L.

In SKIPOGH, CRP levels were measured from blood collected between 2012 and 2016 through standard immunoturbidimetry with different detection limits (1 mg/L or 2 mg/L or 3 mg/L) depending on the centre or diagnostic machine used (Cobas 8000 Roche Diagnostics, Modular Roche Diagnostics, Beckmann). Inter-assay coefficients of variation were 7.5% at 10 mg/L for N=317 (Beckmann), 3.3% at 1.5 mg/L for N=372 (Modular Roche), and 4.0% at 0.9 mg/L for N=344 (Cobas 8000 Roche). Since ∼60% of SKIPOGH participants had CRP values below the detection limits, we imputed left censored values via censored maximum likelihood multiple imputation^40^. We ran 30 imputed data sets with a model of CRP including paternal occupational position (or parental education), body mass index, age, sex, and centre as predictors. Parameters (effects described in section *Mediation approach*) were estimated for each data set of imputed CRPs and pooled following Rubin’s rule.

Gene transcription and DNA methylation in adulthood were considered as potential mediators and were measured from a mixture of leukocytes cells collected in 2011 for YFS and between 2012 and 2016 for SKIPOGH. Transcriptomic data were available for 1,650 YFS and 723 SKIPOGH participants via Illumina chip and sequencing technologies, respectively (see SI Appendix). DNA methylation was measured in 1,548 YFS and 701 SKIPOGH participants via Illumina Infinium chips (see SI Appendix).

Age, sex, centre of blood collection were included as potential confounders.

In main analyses, we excluded individuals with unknown parental occupational position (N=27 [1.6%] in YFS and N=15 [2%] in SKIPOGH), father not working for health reasons (N=1 in SKIPOGH) and missing CRP (N=1 in SKIPOGH) for a total of 1,623 YFS and 706 SKIPOGH participants (mediation analysis). Parental occupational position (or parental education) was modelled as ordered variable, hypothesizing a dose-response relationship^41^. CRP was modelled as a continuous outcome and transformed via the natural logarithm to symmetrize its distribution. Finally, parental occupational position, DNA methylation and transcriptomic data were available in 1,367 YFS and 661 SKIPOGH participants (Bayesian network scoring analysis).

### Pro-inflammatory genes selection

We selected pro-inflammatory genes based on two strategies independently of the study populations. First, we selected genes based on a transcriptome-wide causal inference analysis via 2-sample Mendelian Randomization (2sMR) aiming at establishing leukocytes transcripts (exposure) driving levels of CRP in blood (outcome)^21^. 2sMR is potentially robust with respect to unmeasured confounders and exploits publicly available summary statistics of large-scale GWAS^42^. In our study, we used summary statistics of the associations between about 10M genetic instruments (Single Nucleotide Polymorphisms, SNPs) and gene transcription in leukocytes or CRP levels in blood obtained via GWAS conducted by the eQTLGen consortium^23^ (N=31,684) and the CHARGE consortium^3^ (N=148,164), respectively (see SI Appendix).

Critical to 2sMR are assumptions about validity of genetic variants as instrumental variables^43^. To satisfy the assumption that chosen variants are predictive of the exposure, we selected only eQTLs (*P* value<1.9*10^−5^ corresponding to a false discovery rate <0.05)^23^ and – to reduce bias due to reverse causation – discarded those with a strong association to CRP (*P* value<1.9*10^−5^). Remaining assumptions were addressed by using *cis* eQTLs (likely direct effect of SNP on nearby gene transcription) and applying three complementary 2sMR methods^43^ (see SI Appendix). All 2sMR methods were run with a minimum of 10 instruments. We estimated *F*-statistics to assess instruments strengths (*F*-stat>10 is suggestive of strong instruments). We computed the *q* value^44^ from the *P* values estimated via the three 2sMR methods and declared a gene transcription to be driving CRP levels when *q*≤0.05 in all three 2sMR methods.

Alternatively, we defined our set of pro-inflammatory genes as those having been identified by the conserved transcriptional response to adversity (CTRA) model^12^. The CTRA is composed by 53 genes characterized by up/down-regulation depending on whether their immune function is inflammation- or antiviral/antibody-related, respectively^14^. This gene set has been associated to both early life and adult socioeconomic conditions^12,14^.

### Functional enrichment

We run gene ontology (GO) and pathways enrichment analysis of the list of genes selected via 2sMR with g:Profiler^45^. The list of genes tested via 2sMR was submitted as the background gene list and multiple testing correction was performed with the g:SCS algorithm that takes into account the structure of functionally annotated gene sets^45^. GOs or pathways were declared as significantly enriched when g:SCS adjusted *P* values were <0.05.

### Mediation approach

We adopted a counterfactual mediation framework based on natural effect models^46^ to estimate the portion of the effect of early life socioeconomic conditions on adulthood inflammation transmitted simultaneously or *en-bloc* by transcription levels of multiple genes in leukocytes. Importantly, the identification of these joint indirect effects is not dependent on the true causal order of the mediators^46,47^. This property is useful in our case since the interrelations between the transcription of different genes cannot be determined unequivocally. The magnitude of total and joint indirect effects corresponded to exponentiated parameters of natural effects models and is interpretable as percentage of change in the geometric mean of CRP. The proportion mediated (PM) by a bloc of genes was derived by the ratio of the joint indirect and total effect parameters. See SI Appendix for detailed information on interpretation and estimation of natural effect models’ parameters.

We applied principal component analysis (PCA)^48^ to the selected pro-inflammatory gene sets for estimating a representation of genes’ transcription. In our mediation model, using principal components of genes’ transcription instead of measured transcription levels has the advantage of increasing the reliability of joint indirect effect estimates since it allows to both reduce the number of mediators and the impact of measurement errors in the mediators^49^. Indeed, PCA is an established dimensionality reduction method allowing the data to be described using a small number of uncorrelated variables (the principal components, PCs) while retaining as much information (variance in the original data) as possible. Furthermore, given the ordering of PCs according to decreasing values of explained variance, PCA is also a noise reduction method when retaining only the principal components with highest explained variance^48^.

In main analyses, we selected the number of principal components (PCs) in order to explain about 50% of transcription variance. Confidence intervals (95%) of total and joint indirect effects were estimated through percentiles from 5,000 bootstrap draws (with replacement).

### Local DNA methylation

For each of the pro-inflammatory genes selected either via 2sMR or the CTRA model, we considered DNA methylation sites (CpGs) within 1,500 bp of the annotated gene location. So defined *cis* CpGs should ensure that their potential effect on transcription is direct and not through other genes. Furthermore, we removed cross-reactive CpGs and CpGs with known SNPs in probes having MAF>0.01 to avoid potentially spurious or genetically driven signals.

### Bayesian network model selection

We applied Bayesian network scoring^22^ to select the most likely causal pathway between local DNA methylation and gene transcription that are affected by early life socioeconomic conditions. For each CpG we evaluated the DAG structures represented in Figure S1. For each DAG, a posterior probability was estimated through the Bayesian Gaussian score and uniform priors across the eleven DAGs (*bnlearn* R package^50^). Finally, to alleviate the potential brittleness of inference from a large number of causal structures, we assessed posterior probabilities of 5 groups of DAGs, by specifying a partition of the DAGs^51^. Namely, group A encapsulates models where paternal occupational position does not influence DNA methylation nor transcription levels; group B encapsulates models where paternal occupational position causes changes in DNA methylation and transcription levels, and DNA methylation also causes changes in transcription; group C encapsulates models where paternal occupational position drives transcription levels, but methylation and transcription levels are not related; group D encapsulates models where paternal occupational position drives transcription levels that in turn influences methylation levels; group E encapsulates models where paternal occupational position influences DNA methylation but not transcription levels (see Figure S1). To select the group of DAGs with substantial evidence, we computed a Bayes factor (BF) from the two highest posterior probabilities and choose the group with the natural logarithm of BF larger than 1. In other words, a group of DAGs is selected only if its posterior probability is at least roughly three times larger than any other group’s posterior probability^52^.

### Sensitivity analyses

We investigated the stability of analyses upon removing participants with CRP>10mg/L, which may reflect short term immune responses. We computed indirect effects when summarizing transcription levels with 30% and 60% of explained variance by principal components to assess whether the effect sizes reported in main analyses underestimate the amount of mediation. In SKIPOGH, we run mediation analysis with a binary outcome (CRP≥3 mg/L) as the so defined heightened inflammation was independent of the imputation procedure.

We ran mediation and Bayesian scoring analyses when parental education was used as an indicator of early life socioeconomic conditions. Finally, we repeated the model selection via Bayesian scoring when adding a group of negative control DAGs, namely models where uncoupled DNA methylation and gene transcription drive (both or either one) paternal occupational position.

## Data Availability

Summary statistics for eQTLs were downloaded from https://eqtlgen.org/. Summary statistics for CRP are available upon request to the CHARGE consortium.
SKIPOGH and YFS individual data are available from the PIs of the studies upon reasonable request.

## DATA AVAILABILITY

Summary statistics for eQTLs were downloaded from https://eqtlgen.org/. Summary statistics for CRP are available upon request to the CHARGE consortium.

YFS and SKIPOGH individual data are available from the PIs of the study upon reasonable request.

## Author contributions

S.S. designed the study; M.B., N.D., B.P., M.P., M.Kä., T.L. and O.R. collected the data; C.C., Z.K., P.M., E.P., C.D., O.D., M.K.-I., G.E., M.C.-H., P.V., M.Ki., E.D. and N.V. contributed reagents, materials or analysis tools; C.C. analysed the data; C.C. and S.S. discussed the results and wrote the paper. All authors reviewed and commented on the manuscript.

## Acknowledgements

This work was supported by the European Commission [grant Horizon 2020 number 633666] and the Swiss State Secretariat for Education, Research and Innovation SERI, and by an Ambizione Grant from the Swiss National Science Foundation (PZ00P3_147998).

The YFS study has been financially supported by the Academy of Finland: grants 322098, 286284, 134309 (Eye), 126925, 121584, 124282, 129378 (Salve), 117787 (Gendi), and 41071 (Skidi); the Social Insurance Institution of Finland; Competitive State Research Financing of the Expert Responsibility area of Kuopio, Tampere and Turku University Hospitals (grant X51001); Juho Vainio Foundation; Paavo Nurmi Foundation; Finnish Foundation for Cardiovascular Research ; Finnish Cultural Foundation; The Sigrid Juselius Foundation; Tampere Tuberculosis Foundation; Emil Aaltonen Foundation; Yrjö Jahnsson Foundation; Signe and Ane Gyllenberg Foundation; Diabetes Research Foundation of Finnish Diabetes Association; EU Horizon 2020 (grant 755320 for TAXINOMISIS); European Research Council (grant 742927 for MULTIEPIGEN project); and Tampere University Hospital Supporting Foundation.

The SKIPOGH study was supported by a grant from the Swiss National Science Foundation [grant number 33CM30-124087].

M Kivimäki is supported by the UK Medical Research Council (R024227, S011676), US National Institute on Aging (NIH, R01AG056477), NordForsk and the Academy of Finland (311492). Z Kutalik was supported by the Swiss National Science Foundation (31003A_169929).

The funding sources had no involvement in the study design, data collection, analysis and interpretation, writing of the report, or decision to submit the article for publication.

We are grateful to the CHARGE consortium for sharing their association summary statistics.

## REFERENCES

1. J. Danesh, et al., C-reactive protein and other circulating markers of inflammation in the prediction of coronary heart disease. N Engl J Med 350, 1387–97 (2004).

2. A. Dehghan, et al., Genetic variation, C-reactive protein levels, and incidence of diabetes. Diabetes 56, 872–878 (2007).

3. S. Ligthart, et al., Genome Analyses of > 200,000 Individuals identify 58 loci for chronic inflammation and highlight pathways that link inflammation and complex disorders. Am J Hum Genet 103, 691–706 (2018).

4. R.A. Pollitt, et al., Early-life and adult socioeconomic status and inflammatory risk markers in adulthood. Eur J Epidemiol 22, 55–66 (2007).

5. F. Tabassum, et al., Effects of socioeconomic position on inflammatory and hemostatic markers: a life-course analysis in the 1958 British birth cohort. Am J Epidemiol 167, 1332–1341 (2008).

6. E. Berger, et al., Multi-cohort study identifies social determinants of systemic inflammation over the life course. Nat Commun 10, 773 (2019).

7. R. Castagné, et al., A life course approach to explore the biological embedding of socioeconomic position and social mobility through circulating inflammatory biomarkers. Sci Rep 6, 25170 (2016).

8. C. Carmeli, et al., Mechanisms of life-course socioeconomic inequalities in adult systemic inflammation: Findings from two cohort studies. Soc Sci Med 245, 112685 (2020).

9. R.S. Liu, et al., Socioeconomic status in childhood and C reactive protein in adulthood: a systematic review and meta-analysis. J Epidemiol Community Health 71, 817–826 (2017).

10. G.E. Miller, et al., Low early-life social class leaves a biological residue manifested by decreased glucorticoid and increased proinflammatory signalling. Proc Natl Acad Sci USA 106, 14716–14721 (2009).

11. M.R. Irwin, S.W. Cole, Reciprocal regulation of the neural and innate immune systems. Nat Rev Immunol 11, 625–632 (2011).

12. S.W. Cole, Human social genomics. PLoS Genet 10, e1004601 (2014).

13. N.D. Powell, et al., Social stress up-regulates inflammatory gene expression in the leukocyte transcriptome via beta-adrenergic induction of myelopoiesis. Proc Natl Acad Sci USA 110, 16574–16579 (2013).

14. M.E. Levine, E.M. Crimmins, D.R. Weir, S.W. Cole, Contemporaneous social environment and the architecture of late-life gene expression profiles. Am J Epidem 186, 503–509 (2017).

15. J. Tung, et al., Social environment is associated with gene regulatory variation in the rhesus macaque immune system. Proc Natl Acad Sci USA 109, 6490–6495 (2012).

16. N. Snyder-Mackler, et al., Social status alters immune regulation and response to infection in macaques. Science 354, 1041–1045 (2016).

17. S. Stringhini, et al., Life-course socioeconomic status and DNA methylation of genes regulating inflammation. Int J Epidemiol 44, 1320–1330 (2015).

18. T.W. McDade, et al., Genome-wide analysis of DNA methylation in relation to socioeconomic status during development and early adulthood. Am J Phys Anthropol 169, 3–11 (2019).

19. A.J. Lea, et al., Genome-wide quantification of the effects of DNA methylation on human gene regulation. eLIFE 7, e37513 (2018).

20. F. Guida, et al., Dynamics of smoking-induced genome-wide methylation changes with time since smoking cessation. Hum Mol Genet 24, 2349–2359 (2015).

21. P.C. Haycock, et al., Best (but oft-forgotten) practices: the design, analysis, and interpretation of Mendelian randomization studies. Am J Clin Nutr 103, 965–978 (2016).

22. D. Heckerman, D. Geiger, D.M. Chickering, Learning Bayesian networks: the combination of knowledge and statistical data. Machine Learning 20, 197–243 (1995).

23. U. Võsa, et al., Unraveling the polygenic architecture of complex traits using blood eQTL meta-analysis. bioRxiv:447367 (19 October 2018).

24. G. Hemani, K. Tilling, G. Davey Smith, Orienting the causal relationship between imprecisely measured traits using GWAS summary data. PLoS Genetics 13, e1007081 (2017).

25. J. Bowden, G. Davey Smith, P.C. Haycock, S. Burgess, Consistent estimation in Mendelian randomization with some invalid instruments using a weighted median estimator. Genet Epidemiol 40, 304–314 (2016).

26. O.T. Raitakari, et al., Cohort profile: the cardiovascular risk in Young Finns Study. Int J Epidemiol 37, 1220–1226 (2008).

27. H. Alwan, et al., Epidemiology of masked and white-coat hypertension: the family-based SKIPOGH study. PLoS One 9, e92522 (2014).

28. M. Firmann, et al., The CoLaus study: a population-based study to investigate the epidemiology and genetic determinants of cardiovascular risk factors and metabolic syndrome. BMC Cardiovasc Disord 8, 6 (2008).

29. I. Guessous, M. Bochud, J.-M. Theler, J.-M. Gaspoz, A. Pechère-Bertschi, 1999-2009 trends I prevalence, unawareness, treatment and control of hypertension in Geneva, Switzerland. PLoS One 7, e39877 (2012).

30. N.E. Banovich, et al. Methylation QTLs are associated with coordinated changes in transcription factor binding, histone modification, and gene expression levels. PLoS Genet 10, e1004663 (2014).

31. S. Wang, W. Wu, F.X. Claret, Mutual regulation of microRNAs and DNA methylation in human cancers. Epigenetics 12, 187–197 (2017).

32. M. Loi, L. Del Savio, E. Stupka, Social Epigenetics and Equality of Opportunity. Public Health Ethics 6, 142–153 (2013).

33. D. Cano-Rodriguez, M.G. Rots, Epigenetic editing: On the verge of reprogramming gene expression at will. Curr Genet Med Rep 4, 170–179 (2016).

34. D. Schübeler, Function and information content of DNA methylation. Nature 517, 321–326 (2015).

35. M. Gutierrez-Arcelus, et al. Passive and active DNA methylation and the interplay with genetic variation in gene regulation. Elife 2, e00523 (2013).

36. L.T. Husquin, et al. Exploring the genetic basis of human population differences in DNA methylation and their causal impact on immune gene regulation. Genome Biol 19, 222 (2018).

37. B. Galobardes, J. Lynch, G. Davey Smith, Measuring socioeconomic position in health research. Br Med Bull 81-82, 21–37 (2007).

38. D. Rose, E. Harrison, The European socio-economic classification: A new social class schema for comparative European research. Eur Soc 9, 459–490 (2007).

39. S. Stringhini, et al., Socioeconomic status and the 25×25 risk factors as determinants of premature mortality: a multicohort study and meta-analysis of 1.7 million men and women. Lancet 389, 1229–1237 (2017).

40. J. Boss, et al., Estimating outcome-exposure associations when exposure biomarker detection limits vary across batches. Epidemiology 30, 746–755 (2019).

41. P. Vineis, et al. Health inequalities: Embodied evidence across biological layers. Soc Sci Med 246; 112781 (2020).

42. D.A. Lawlor, Commentary: Two-sample Mendelian randomization: opportunities and challenges. Int J Epidemiol 45, 908–915 (2016).

43. J. Bowden, et al., A framework for the investigation of pleiotropy in two-sample summary data Mendelian randomization. Stat Med 36, 1783–1802 (2017).

44. J.D. Storey, The positive false discovery rate: a Bayesian interpretation and the q-value. Ann Statist 31, 2013–2035 (2003).

45. U. Raudvere, et al., g:Profiler: a web server for functional enrichment analysis and conversions of gene lists (2019 update). Nucleic Acids Res 47, W191–W198 (2019).

46. J. Steen, T. Loeys, B. Moerkerke, S. Vansteelandt, Flexible mediation analysis with multiple mediators. Am J Epidemiol 186, 184–193 (2017).

47. T.J. VanderWeele, S. Vansteelandt, Mediation analysis with multiple mediators. Epidemiol Methods 2, 95–115 (2014).

48. I.T. Jolliffe, J. Cadima, Principal component analysis: a review and recent developments. Philos Trans A Math Phys Eng Sci. 374, 20150202 (2016).

49. L. Valeri, X. Lin, T.J. VanderWeele, Mediation analysis when a continuous mediator is measured with error and the outcome follows a generalized linear model. Stat Med 33, 4875–4890 (2014).

50. M. Scutari, J.B. Denis, Bayesian networks with examples in R. (CRC Press, 2015).

51. W.D. Penny, et al., Comparing families of dynamic causal models. PLoS Comput Biol 6, e1000709 (2010).

52. A.E. Raftery, “Bayesian model selection in social research”. In Sociological methodology, P.V. Marsden, Ed. (Blackwell, 1995), pp. 111–196.

